# Integrating HPV vaccination into adolescent health and routine immunization delivery platforms in rural and pastoralist Ethiopia: A mixed-methods implementation study of the HPV-VISION program

**DOI:** 10.64898/2026.09.21.26363592

**Authors:** Rahel D. Gebreyohannes, Claire W. Rothschild, Selamawit Hirpa, Helen Negassi, Abiy S. Estifanos, Damen Hailemariam, Meselech A. Roro, Bekele Belayihun, Susannah Gibbs, Claire E. Moodie, Kasahun Girma, Lidiya Berihun, Biruk Hailu, Seyoum Atlie, Melkamu Ayalew, Nahom Solomon, Muhammedawel Kaso, Wakgari Deressa

## Abstract

**Introduction:** Vaccination against human papillomavirus (HPV) is the most effective strategy for preventing cervical cancer. We evaluated HPV Vaccine Integrated Service Implementation (HPV-VISION), a program aimed at integrating HPV vaccination with adolescent health (AH) services and the Expanded Program on Immunization (EPI) to improve vaccine uptake among girls aged 9 to 14 years in rural Ethiopia.

**Methods:** This mixed-methods implementation study was implemented in four woredas testing two models of integrating HPV vaccination into routine health services: integration with EPI and with an AH service package. We used program monitoring data; exit interviews, in-depth interviews, and focus group discussions with adolescent girls and caregivers; key informant interviews; and costing data. We analyzed effectiveness using negative binomial generalized estimating equation models with facility-level monthly HPV vaccinations as the primary outcome. We analyzed exit interview data descriptively across models. We analyzed qualitative data thematically and conducted a costing analysis.

**Results:** The HPV-VISION program delivered 25,821 HPV vaccinations from March to August 2025. Adjusting for baseline differences, facilities in the AH model delivered, on average, 34% fewer HPV vaccinations per month compared to those in the EPI model (adjusted IRR: 0.66; 95% CI: 0.51-0.87). Nearly all girls said they would be happy to receive services again at the place they were interviewed (EPI: 94%, AH: 98%). Experience of care was positive overall with individual measures varying across models. The incremental delivery cost (excluding vaccine cost) per vaccinated girl was USD $7.54 in the EPI model and USD $14.28 in the AH model.

**Conclusion:** We recommend that HPV vaccine delivery incorporates integration with AH services and routine vaccination to most efficiently reach underserved girls. An optimal mix of integration with the EPI program and AH services should be based on regional HPV vaccination indicators, other AH needs, population demographics, and costs of the models.

**KEY MESSAGES:** *What is already known on this topic:* Globally and in Ethiopia, HPV vaccine delivery relies heavily on school-based platforms and targeted outreach initiatives to reach large cohorts of eligible adolescents Addressing inequities in HPV vaccination coverage will require complementary facility-based and community-based strategies that directly engage girls while also addressing cultural misconceptions, mistrust, and other social barriers.

*What this study adds:* We found that two models for integrating HPV vaccination into routine healthcare delivery for girls aged 9 to 14 in Ethiopia performed well and were highly acceptable. Qualitative research confirmed perceptions that community engagement, school involvement, and health facility linkages were central to successful implementation, while exit interview data indicated that clients served by both models had positive experiences and were satisfied with the care they received.

*How this study might affect research, practice or policy:* We recommend that Ethiopia’s strategy to scale-up integrated HPV vaccine delivery maintains the existing lower cost school-based campaign approach, while also incorporating integration with both routine immunization and adolescent health services to most efficiently reach adolescent girls who are seeking various services, including puberty education and menstrual health products and services.

## BACKGROUND

In sub-Saharan Africa, cervical cancer incidence has been rising across many countries over the past two decades [1]. It accounts for over 80% of the global burden of the disease, ranking as the second most prevalent cancer among women and the leading cause of cancer-related mortality in this population [2]. Vaccinating 9-to 14-year-old girls for the human papillomavirus (HPV) is highly effective and cost-effective, reducing the burden of cervical cancer in women by up to 90% when administered before the onset of sexual activity [3]. The World Health Organization (WHO) has set a global target of vaccinating 90% of girls by age 15 as part of its strategy to eliminate cervical cancer in women by 2030 [4]. However, vaccine uptake among girls aged 9 to 14 years in many low- and middle-income countries (LMICs), particularly in Africa, remains suboptimal.

In Ethiopia, cervical cancer is the second most common cancer among women after breast cancer and is estimated to cause 7,500 new cases and about 5,300 deaths annually [5]. Ethiopia introduced the HPV vaccine into its immunization program in 2018 in a single-age cohort of 14-year-old girls through school-based campaigns [6]. The first large-scale campaign successfully vaccinated over 1.5 million girls. With increased availability of HPV vaccines, Ethiopia updated its national HPV guidelines in 2021 to target a multi-age cohort (MAC) of girls ages 9 to 14 with a two-dose schedule [6] and in 2024 switched to a single dose schedule following WHO guidance [7,8]. Despite efforts to expand coverage, HPV vaccination among girls in Ethiopia remains suboptimal. A recent meta-analysis estimated the national average uptake among schoolgirls at only 42% [9]. Population-based evidence is limited, with uptake reported at around 50% in various parts of the country [10,11], far short of the global target of vaccinating 90% of girls by the age of 15 to eliminate cervical cancer by 2030 [4].

Globally, HPV vaccine delivery relies heavily on school-based platforms and targeted outreach initiatives to reach large cohorts of eligible adolescents [12]. In Ethiopia, this pattern is mirrored, as HPV vaccination is delivered almost exclusively through periodic school-based campaigns with limited use of health facility or community-based outreach. While school-based strategies prove convenient and effective for increasing HPV vaccination coverage, they face logistical challenges, parental consent issues, and exclusion of out-of-school girls [13,14]. Ethiopia’s reliance on school-based campaigns leaves many rural and out-of-school girls unreached. These girls face intersecting barriers, including school drop-out, limited mobility and access to health information, and restrictive social norms that promote early marriage and childbearing [15,16].

Addressing inequities in HPV vaccination coverage will require complementary facility-based and community-based strategies that directly engage girls while also addressing cultural misconceptions, mistrust, and other social barriers [15]. Integrating vaccine delivery with routine health services, including those designed specifically for adolescents, offers potential to expand access and improve coverage among targeted girls using existing health systems [13,17]. Previous studies have emphasized the need for comprehensive assessments that consider not only costs but also the practical challenges of vaccine delivery, program sustainability, and strategies to improve access among hard-to-reach and out-of-school girls [18]. The goal of this implementation research study was to assess and compare two approaches to HPV vaccine delivery: integration of HPV vaccination into the Expanded Program on Immunization (EPI) routine vaccination program and integration of HPV vaccination with adolescent health (AH) services. The aims were to evaluate the effectiveness of HPV vaccine delivery for girls ages 9 to 14 and to assess the feasibility, acceptability, and costs for the two delivery approaches.

## METHODS

### Study design

This quasi-experimental, mixed-methods approach evaluated integrated HPV vaccine delivery models among girls ages 9 to 14 years in four rural intervention woredas in three purposively selected regions of Ethiopia (Oromia, Sidama, and Somali). These sites were selected to represent diverse geographical and demographic contexts. Selection criteria for woredas included a large population of 9-to-14-year-old girls, active participation in Smart Start, a substantial out-of-school population, and relatively low baseline HPV vaccination coverage (among candidate sites). Four additional woredas from these regions were selected as comparison areas where the integrated models were not implemented using the same selection parameters. With the exception of service statistics captured through program monitoring and evaluation (M&E) systems, no primary data was collected from these areas, as formative research indicated no ongoing HPV vaccination activities at the time of the study. The intervention woredas were prospectively non-randomly assigned to two implementation arms (described in detail below). The EPI arm was implemented in Gomma and Awbare woredas, and the AH arm was implemented in Sokoru and Hawella woredas. Awbare woreda in the Somali region was selected to include a pastoralist community, while the other woredas represent rural agrarian populations.

The Ethiopian Public Health Association (EPHA) Institutional Review Board (IRB), the PSI Research Ethics Board, and the Heartland IRB all approved the study. Written informed consent was obtained from all adult participants, and parental consent was obtained for girls accompanied by a parent or guardian. Girls aged 9 to 14 years old provided verbal assent; a waiver of parental consent was obtained for unaccompanied girls ages 13 to 14. The trial was prospectively registered on ClinicalTrials.gov (NCT06667323).

### Patient and public involvement statement

Patients and members of the public were not involved in the development of the research objectives, study design, outcome measures, or data collection instruments. However, community members residing in the study areas were involved through three mechanisms: first, community members were recruited to participate in intervention co-design workshops, which occurred during the formative phase and engaged participants in interpreting the formative phase study results to finalize the intervention design; community members were also engaged after approximately 6 months of implementation in “pause-and-reflect” workshops to review study findings from the first implementation phase to refine the implementation models. A community advisory board comprised of religious leaders, healthcare providers, community mobilizers, and school directors was also convened bi-monthly to monitor implementation progress. The engagement of members of the public was therefore focused on design and iterative refinement of the intervention being studied, rather than design of the research study specifically.

### Vaccine delivery models

Smart Start is an initiative that uses financial planning as a gateway to engage young married couples in discussions about delaying first births, spacing pregnancies, and using modern contraception to achieve life goals. Scaled nationally within the Ministry of Health’s Health Extension Program (HEP), it now reaches over one million rural adolescent girls and young couples in Ethiopia. The integrated models being tested in this study each leveraged Smart Start’s community-based demand generation and mobilization structures, including health extension workers (HEWs), women’s development army members (WDAs), traditional birth attendants (TBAs), village health leaders (VHLs), and household-level outreach systems, to identify and refer eligible girls for vaccination. HPV-related data management was integrated into Smart Start’s DHIS2-based M&E system, enabling streamlined monitoring of HPV vaccination demand generation activities and service delivery. A detailed description of activities is provided in supplemental material (Figure S1).

The HPV-VISION program tested two HPV vaccination delivery models, with the study including a third standard-of-care arm for comparison. The delivery models were informed by a formative phase, during which key informant interviews, facility-readiness assessments, and four co-design workshops (one national- and three woreda-level) were conducted to design the detailed implementation package for each model.

#### Comparison arm

In the comparison arm, no HPV-VISION activities other than routine M&E were implemented. The selected comparison areas were expected to implement standard-of-care HPV vaccine delivery, which – at the time of initial study design – was anticipated to be school-based, annual MAC campaigns that made up the existing national strategy for HPV vaccination delivery. However, no school-based campaigns were conducted during HPV-VISION implementation.

#### EPI arm

In the EPI arm, HPV vaccination was delivered through Ethiopia’s existing routine immunization system. The model focused on strengthening the supply side of EPI delivery by ensuring uninterrupted HPV vaccine availability at both health centers and posts, reinforcing cold-chain management, training health workers on HPV vaccine administration and documentation, and improving coordination between immunization and primary healthcare teams. Demand generation was conducted through a woreda-specific combination of HEWs, WDAs, TBAs, and VHLs who implemented targeted household mobilization to identify eligible girls – particularly those missed by school-based campaigns – and refer them for vaccination at health posts, health centers, or outreach sites.

#### Adolescent health services arm

The AH model included the supply- and demand-side elements of the EPI model, including demand creation and supply-side activities to ensure vaccine supply and service availability. However, in addition to receiving HPV vaccination, girls were offered AH services such as puberty and menstrual health counselling, menstrual pads, general health counselling, and nutrition screening services. As in the EPI arm, HEWs, WDAs, VHLs, and TBAs conducted demand generation and community mobilization tailored to both HPV vaccine awareness and broader adolescent wellbeing, while HEWs at health posts and healthcare providers at health centers delivered both the vaccine and AH services during routine and outreach sessions. At the health post, AH services were typically conducted as a fully integrated package – provided in the same space by the same HEW. In health centers, however, intra-facility referral was required between AH services, typically provided in the family planning or youth friendly service room, and HPV vaccination, which was provided in the EPI unit of the building.

All sites implementing the EPI and AH models received ongoing support throughout implementation, including routine supportive supervision visits and performance review meetings. All sites, including those in the comparison arm, received standard M&E support.

### Effectiveness analysis

Data were extracted from routine program M&E data, in which HPV vaccinations (total, by age, and by current school enrollment status) were recorded using paper-based forms at each health center and post supported by the intervention. The primary outcome was the monthly number of HPV vaccinations delivered per facility to 9-to 14-year-olds. Secondary outcomes included monthly HPV vaccinations per facility to out-of-school girls, in-school girls, and by year of age.

We fit a population-averaged negative binomial generalized estimating equation (GEE) model to assess associations between intervention arm (AH vs. EPI) and the primary and secondary outcomes. The primary model included a variable for intervention arm as the primary exposure and the following adjustment variables: month of implementation; facility type (health center vs. health post); in-school eligible girls and out-of-school eligible girls mapped for the November 2024 MAC campaign; number of in-school and out-of-school girls vaccinated during the MAC campaign; and number of in-school and out-of-school unvaccinated girls mapped by the HPV-VISION program in March 2025.We fit analogous fully adjusted GEE models for the secondary outcomes. All models used an exchangeable working correlation structure with robust standard errors to account for repeated measures at the facility level. For visualization purposes only, the primary GEE model was additionally fitted with interaction terms between month and intervention arm. Models included data from April through August 2025, which represents full-intensity implementation across all intervention areas.

Missing data for monthly vaccine delivery were treated as zero because they are recorded as missing in the M&E system when no vaccine delivery occurs. Because MAC mapping, vaccination performance, and program mapping were not consistently recorded at health centers, we coded these as zero for all health centers to allow this information to contribute to the model for health posts. We then tested effect modification by facility type. Finally, we conducted additional sensitivity analyses to examine the possible effect of a national polio and measles outreach campaign into which HPV vaccination was integrated in one of the intervention woredas in May 2025.

### Exit interviews

A convenience sample of caregiver/girl (ages 9-14) dyads was surveyed separately after receiving services at participating facilities in intervention woredas. For the precision-based sample size calculation, we conservatively assumed 50% prevalence of outcomes of interest (e.g., person-centeredness of care) to yield 97 caregiver-child dyads per intervention woreda (388 total each for caregivers and girls 9–14), producing pooled estimates with ±5% margin of error with α=0.05 (10%-point width), rounded up to 400 per group. The approved protocol permitted increasing the sample size if feasible to improve precision at the woreda level. Among unaccompanied girls, the age range was restricted to 13 to 14 years. Caregivers were included if they were the parent or legal guardian of an eligible girl and were 18 years or older. Participants were excluded if they required urgent medical treatment or referral at the time of recruitment. Girls who did not receive an HPV vaccination on the day of the interview were excluded from the analysis (along with their caregivers). Exit interviews were conducted using a structured tool that captured socio-demographic characteristics, HPV vaccination attitudes and beliefs, healthcare utilization, quality of care, satisfaction, and experiences with AH services. The tool was initially developed in English based on relevant literature and local context, then translated into local languages (Amharic, Afan Oromo, Sidama, and Somali), and pretested.

Exit interviews were conducted in May and June 2025. Data were collected electronically using the Open Data Kit (ODK) platform on smartphones and tablets in English and local languages. Healthcare workers linked eligible girls to study staff immediately after girls received services. Eligibility was assessed by research assistants using a short screening instrument before obtaining informed consent or assent. Interviews.

Exit interview data were analyzed using descriptive statistical methods. Data cleaning and analysis were done using SPSS version 26. Descriptive analysis and tabulations were used to summarize all variables, including frequency distributions for categorical variables and computation of means, medians, and standard deviations for continuous variables. Dichotomous and categorical variables were compared across intervention models using unadjusted regression models appropriate for each dependent variable type (logistic, ordinal logistic, and multinomial logistic regression), with standard errors clustered at the health facility in which the interview was conducted.

### Qualitative data

In-depth interviews (IDIs) (n=42) were conducted with caregivers and girls ages 9 to 14, while focus group discussions (FGDs) included four with caregivers of both vaccinated and unvaccinated girls and six with girls, representing both in-school/out-of-school groups and vaccinated/unvaccinated groups. Qualitative sample sizes were determined based on thematic saturation. Participants were recruited from intervention woredas based on the eligibility criteria with the assistance of “community guides”, including HEWs, WDAs, TBAs, and VHLs. Smart Start focal people and HEWs helped the research team identify key informants (n=24) with extensive experience in the HPV vaccination program at national, regional, woreda, and community levels. They included Ministry of Health officials, regional health bureau staff, woreda health office personnel, health facility staff, religious and community leaders, WDAs, VHLs, and TBAs engaged in HPV vaccination activities.

Semi-structured interview guides were developed in English and refined based on input from the study research team, translated into four local languages, and back-translated to ensure accuracy and consistency. The guides were designed using the Reach, Effectiveness, Adoption, Implementation, and Maintenance (RE-AIM) framework to explore implementation experiences. Key informant interviews (KIIs) focused on identifying effective implementation strategies. IDIs and FGDs assessed acceptability, access, and experiences with AH services and HPV vaccination. Participatory techniques, including vignettes, ranking, and card sorting, were used with adolescent girls to facilitate age-appropriate engagement and explore perceptions and social norms related to gender and health behaviors.

Data were collected face-to-face in private settings in July 2025. All interviews and FGDs were audio-recorded and supported by detailed field notes. Qualitative data were transcribed and analyzed thematically using a hybrid deductive-inductive coding approach. We identified themes related to implementation, perceived effectiveness, feasibility, acceptability, and scalability. Quotes from participants were incorporated to substantiate key themes, maintain contextual meaning, and enhance credibility of the results.

### Costing analysis

Costs were captured in a subsample of implementation facilities in Gomma (to represent the EPI model), Sokoru (AH model), and Teticha (comparison arm); because there was no HPV vaccination during the study period, comparison arm sites were used to capture costs of the most recent MAC campaign. For AH and EPI sites, high performing facilities were purposively selected and therefore represent optimized implementation conditions rather than average performance. We applied a four-step approach: (1) identifying all expenses related to integrating HPV vaccination into existing services; (2) quantification of each expense using standard market prices and salary scales; (3) comparing costs across delivery models to estimate incremental costs; and (4) calculating costs per vaccinated girl. MAC costs for the 2024 campaign were assessed by interviewing key informants from the central government and from health centers and health posts. Personnel costs were estimated based on employees’ gross earnings. Capital costs for equipment, vehicles, and buildings were calculated using replacement costs and estimated useful life, while construction costs were valued per unit area. Both capital costs and construction costs were allocated to HPV vaccination services based on the proportion used for HPV vaccination. The incremental cost of each HPV vaccination strategy was determined by dividing the total additional cost incurred by the total number of girls vaccinated in the sampled sites for the period of March through July 2025.

## RESULTS

### Effectiveness

From March to August 2025, the HPV-VISION program supported delivery of 25,821 HPV vaccinations within 157 public health centers and posts. Overall, 17,985 of vaccinations were delivered through 93 sites (16 health centers and 77 health posts) implementing the EPI model, with a mean total HPV vaccination count of 193.4 per facility. The remaining 30% of HPV vaccinations (7836/25,821) were delivered through 63 sites implementing the AH model, with a per-facility mean of 124.4 HPV vaccinations (Table S1). Nearly two-thirds (63%) of HPV vaccinations were delivered to out-of-school girls, varying from 70% in the EPI model to 48% in the AH model. Holding covariates constant, including the number of eligible girls per facility, facilities in the AH model delivered 34% fewer HPV vaccinations per month than those in the EPI model (adjusted IRR [aIRR]: 0.66, 95% confidence interval [CI]: 0.51-0.87) (Figure 1, Table 1).

**Table 1.**
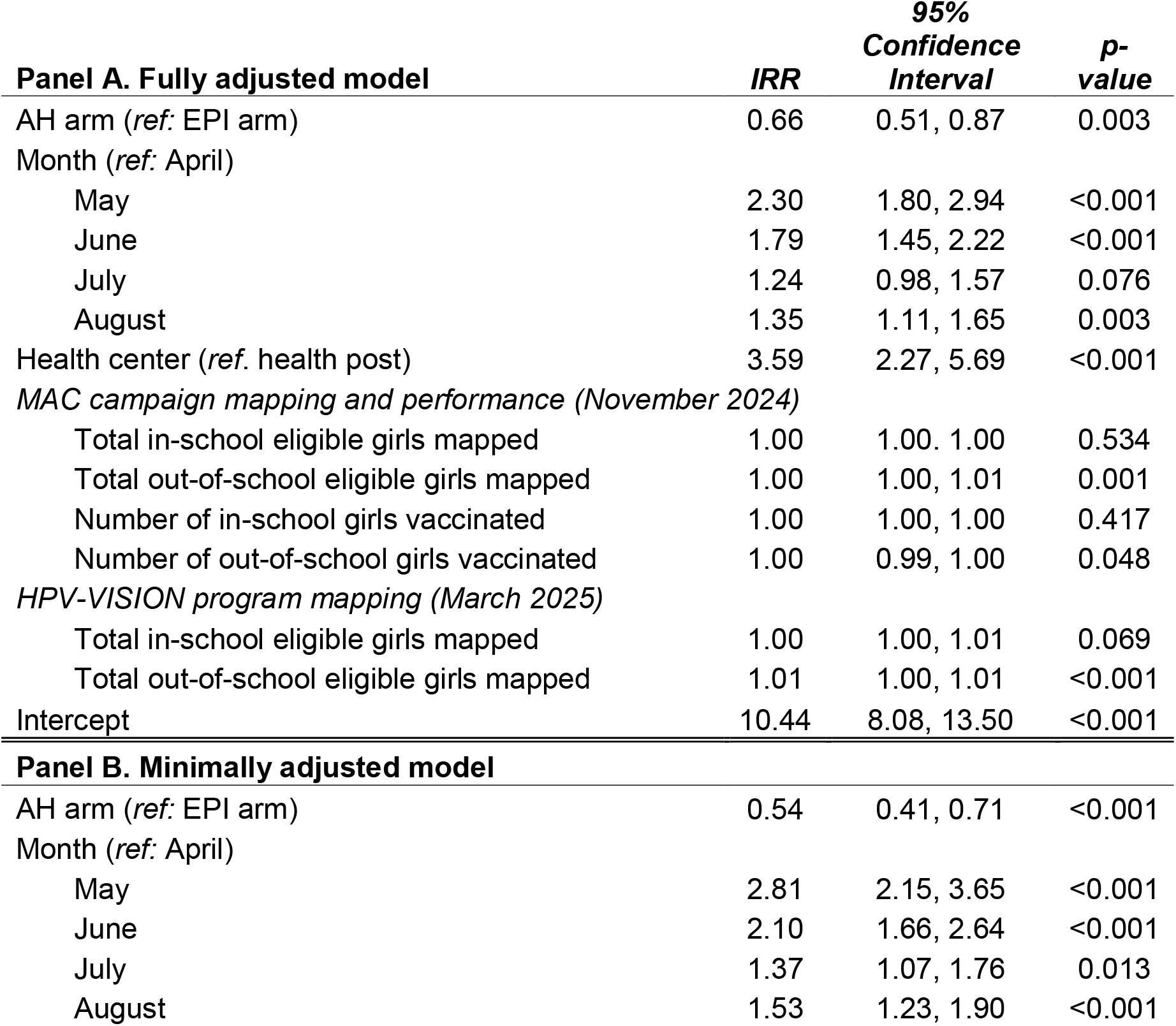
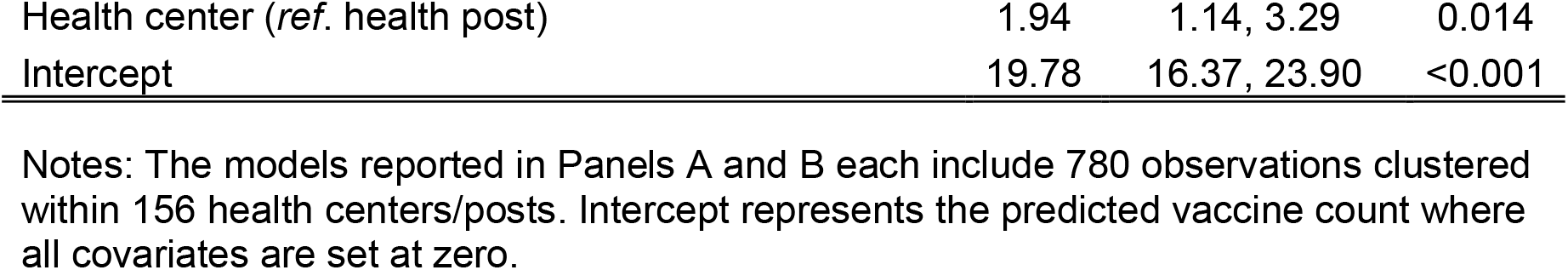
Negative binomial GEE regression results from primary outcome model.

**Figure 1.**
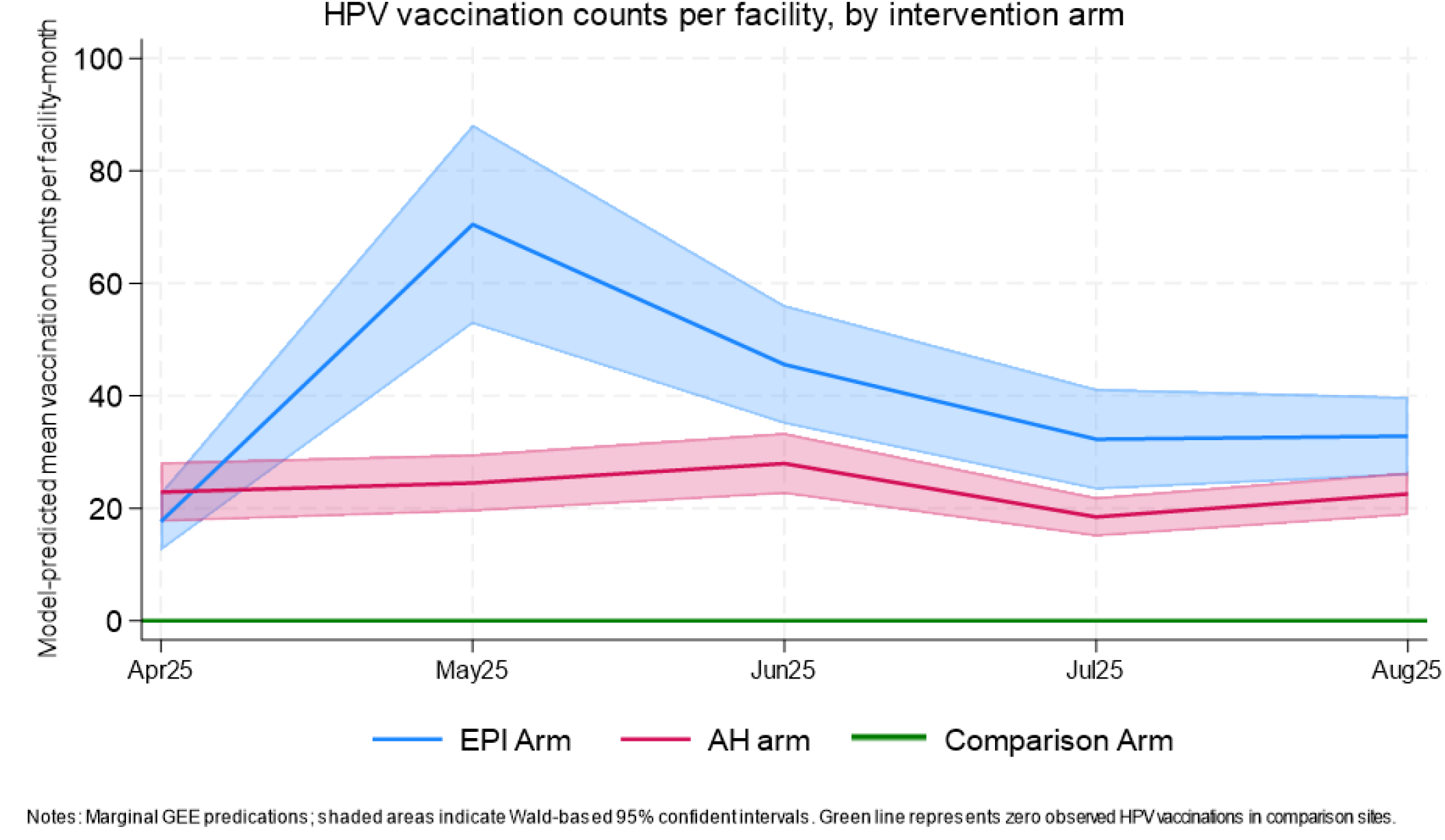
Predicted HPV vaccination counts per facility, by intervention arm. Note: 780 month-level observations within 156 health centers/posts

We found similar associations between intervention arm and most secondary outcomes (Figure S2, Table S2). However, we found no association between intervention arm and HPV vaccination for in-school girls or for 13-or 14-year-olds. Facility type modified the association between intervention arm and total HPV vaccination: compared with health posts, health centers showed a larger relative reduction in HPV vaccinations under the AH arm compared with the EPI arm (P=0.004) (Figure S3, Table S3 & S4) with no difference in HPV vaccinations between the AH and EPI arms within health posts (P=0.24). Models excluding observations from Awbare woreda and all observations from the month of May, to account for the measles campaign, were slightly attenuated and no longer statistically significant (Table S2).

These effectiveness results align with data from qualitative interviews with key informants. In both arms, key informants reported that HPV vaccine uptake increased during the implementation period of this project, especially for out-of-school girls:

> *“In the past months, we identified those in school and out of school*… *As a result, uptake of this vaccine increased…” [WDA, AH arm]*
>
> *“…In these three months, except the first month, we overachieved our targets in the other two. This shows that we conducted a lot of awareness activities to reach this milestone [Increased number of HPV vaccinated girls]” [Healthcare provider, EPI arm]*

### Feasibility

WDAs, VHLs, and TBAs actively identified and mapped eligible girls, including those in remote areas, and linked them to health facilities for vaccination in both the EPI and AH models. Key informants highlighted that identifying, mapping, and linking eligible girls was feasible, and routinizing the HPV vaccine was beneficial for reducing service delivery resources compared to campaign-based programs.

> *“Now, in the third month of implementation, we are also reaching places that are far from health services, such as hard-to-reach areas. We mobilize girls and link them to health facilities. We also work with teams such as VHLs and TBAs. These community members visit villages, identify girls aged 9 to 14 who have not received the vaccine, register them, and bring them to health facilities so that we can vaccinate them*.*” [Healthcare provider, EPI arm]*

Moreover, as discussed by several key informants, routine integration of HPV vaccination required fewer resources compared to the campaign-based approach.

> *“There is a big difference in providing this service in the form of a campaign and in the routine. When you provide this service through a campaign, you may require a significant number of resources. However, if it is a routine, it can be provided at the existing health facility by health professionals without any cost or expenditure*.*” [MCH director from the Regional Health Bureau]*
>
> *“There is no other special resource that’s necessary. We have already selected the people. It’s easy to cascade it…” [EPI focal person, AH arm]*

### Acceptability and client experience

Qualitative data across both study arms indicated that integrated HPV vaccination delivery was acceptable to healthcare providers and community members. In the Sokoru and Hawella districts, integration of HPV vaccination with AH services was reported to be acceptable. The provision of reusable menstrual pads encouraged adolescent girls to visit health facilities for vaccination. Health providers also offered counselling on menstrual health, nutrition, and other AH topics, which further motivated participation.

> *“There is a change. Most girls have now started protecting themselves from this disease. They come to the health post to ask for the vaccine. Especially the pads we were supplied with became very helpful, and girls come and take the vaccine*…*” [HEW, AH arm]*
>
> *“In my opinion, it must be delivered at health posts or health centers…That is why I believe providing the service at health facilities is better than at schools…As I mentioned above, if the vaccination is provided at school [as the previous campaign period], the girl may feel uncomfortable receiving the services. However, if it is offered at health facilities, it is more appropriate for adolescent girls to take advantage of the services there*.*” [Father with unvaccinated girl, AH arm]*
>
> *“The routine vaccination is always available. Before, it was delivered through campaigns, and if someone missed it today, they wouldn’t get another chance. But now, it’s accessible at any time. That’s the good thing about routine vaccination*.*” [HEW, EPI arm]*

These findings were substantiated in client exit interviews (Table 2). Most caregivers across both models reported positive experiences of care (Table 3). Four of the five experience measures were higher in the EPI arm (*P*-values all <.01), whereas the measure of shared decision making was similar in the AH arm (EPI: 60%, AH: 71%, *P*=.23). Very few girls were concerned that someone might judge them for receiving services (EPI: 10%, AH: 3%, *P*=.02), and nearly all girls said they would return to the place they received services for care in the future (EPI: 94%, AH: 98%, P=.24).

**Table 2.** Background characteristics of girls receiving HPV vaccination on the date of interview and their caregivers Caregivers (n=374)

| Variable | EPI model | AH model | $P$-value |
| --- | --- | --- | --- |
| **Girls (n=416)** |  |  |  |
| Age (N=416) |  |  | 0.53 |
| 9-12 years old | 106 (65.8%) | 153 (60.7%) |  |
| 13-14 years old | 55 (34.2%) | 99 (39.3%) |  |
| Educational attainment (N=416) |  |  | 0.48 |
| Never attended | 7 (4.3%) | 11 (4.3%) |  |
| Just started school | 8 (4.9%) | 3 (1.2%) |  |
| Primary level (Grade 1-4) | 112 (69.1%) | 175 (68.9%) |  |
| Primary level/junior secondary (Grade 5-8) | 31 (19.1%) | 64 (25.2%) |  |
| Secondary/high school (Grade 9-12) | 4 (2.5%) | 1 (0.4%) |  |
| Currently enrolled in school (N=398) | 123 (79.4%) | 207 (85.2%) | 0.44 |
| Type of participant (girl) (N=416) |  |  | 0.46 |
| Accompanied | 151 (93.2%) | 223 (87.8%) |  |
| Unaccompanied | 11 (6.8%) | 31 (12.2%) |  |
| Previously heard of HPV vaccine | 91 (56%) | 207 (82%) | 0.001 |
| **Caregivers (n=374)** |  |  |  |
| Region (N=374) |  |  | NA |
| Oromia | 81 (53.6%) | 94 (42.2%) |  |
| Somali | 70 (46.4%) | NA |  |
| Sidama | NA | 129 (57.8%) |  |
| Sex (N=371) |  |  | 0.68 |
| Female | 136 (90.7%) | 195 (88.2%) |  |
| Male | 14 (9.3 %) | 26 (11.8%) |  |
| Currently married (N=371) | 133 (88.7%) | 189 (85.5%) | 0.49 |
| Highest educational status (N=371) |  |  | 0.08 |
| Never attended | 92 (61.3%) | 98 (44.3%) |  |
| Primary level (Grade 1-4) | 19 (12.7%) | 56 (25.3%) |  |
| Junior secondary (Grade 5-8) | 24 (16.0%) | 42 (19.0%) |  |
| High School (Grade 9-12) | 14 (9.3%) | 24 (10.9%) |  |
| Tertiary level (University/college) or higher | 1 (0.7%) | 1 (0.5%) |  |
| Religion (N=371) |  |  | <.001 |
| Muslim | 145 (96.7%) | 97 (43.9%) |  |
| Protestant | 1 (0.7%) | 97 (43.9%) |  |
| Other (Orthodox, Catholic, Hawariyat) | 4 (2.7%) | 28 (11.6%) |  |
| Previously heard of HPV vaccine | 107 (71%) | 184 (83%) | 0.04 |

**Table 3:** Experience of care and satisfaction among caregivers and girls.

| Variables* | EPI model | AH model | P-value |
| --- | --- | --- | --- |
| **Caregivers** |  |  |  |
| Caregiver and child treated with respect most/all of the time | 147 (98.0%) | 175 (81.0%) | 0.001 |
| Health care providers involved us in decisions about girls care most/all of the time | 90 (60.0%) | 152 (71.4) | 0.23 |
| Health workers explained about any examinations/procedures provided to my girl most/all of the time | 129 (86.0%) | 121 (57.6%) | <0.001 |
| I trust health care providers about girls care most/all of the time | 145 (96.7%) | 170 (76.0%) | 0.001 |
| Health care provider paid attention to you and your girl most/all of the time | 137 (91.3%) | 163 (77.6%) | 0.005 |
| **Girls** |  |  |  |
| I am worried that someone will judge me for getting the health services I got today. | 16 (9.9%) | 8 (3.1%) | 0.02 |
| If I needed health services, I would be happy to come back here again in the future. | 152 (93.8%) | 250 (98.4%) | 0.24 |

\*Don't know and non-responses were treated as missing

### Scalability

The integrated models for HPV vaccine delivery demonstrated accessibility, continuity, and sustained vaccine delivery. Key informants in both arms emphasized the need to scale up HPV vaccine integration nationwide.

> *“This [integrated HPV vaccination service] needs to be repeated in every woreda…… I hope it expands to every woreda in Ethiopia. This is saving the life of adolescent girls” [EPI focal person, AH arm]*
>
> *“We expect this vaccine to become part of the routine immunization services provided by health institutions. It has already been initiated [as a pilot project], and what remains is to fully integrate it as a regular service [in every corner of the country] and continue to promote it…” [EPI focal person, EPI arm]*

However, interruptions in the HPV vaccination program across both arms, caused by various factors, posed threats to scalability. Irregular vaccine availability at health posts was a recurring problem, and services were often provided only after specific mobilization efforts. While the program proactively identified and addressed supply-side barriers, including vaccine distribution from central stores and cold chain capacity, both the EPI and AH models rely on the underlying EPI and health extension program (HEP) infrastructure in order to ensure vaccination services are available. This led to frustration among caregivers and community volunteers who brought girls for vaccination.

> *“After I recovered from my illness, the WDA told me to go and take the vaccine, so I went to the health post, but it wasn’t available, and she [HEW] was not there*.*” [Unvaccinated girl, EPI arm]*
>
> *“We mobilize the children based on the HEWs’ guidance. However, if the girls come at unscheduled times, they might not get the vaccination*.*” [WDA, AH arm]*

Additionally, at AH arm sites, participants discussed frequent stockouts of menstrual pads and raised concerns regarding the sustainability of menstrual pad supply. Furthermore, non-functional or poorly established youth-friendly service units were identified as barriers to integrating the HPV vaccination program with AH services.

> *“It is better to provide this service along with sanitary pads; however, sustainability is a concern, as our facility struggles even to obtain some items like medicine, let alone pads*.*” [Immunization expert, Regional Health Bureau]*
>
> *“The Smart Start service, and the HPV vaccine demand creation were supposed to be integrated into the Youth Friendly Service units, but the problem is that the YFS [youth-friendly services] units are not functioning well*.*” [HPV vaccine focal person, Regional Health Office]*

### Costs

Delivery costs were lowest for the MAC model and highest for the AH model (Table 4). The total project cost, excluding vaccine cost of USD 4.60 per vaccinated girl, for delivering the HPV vaccine through the MAC campaign strategy in Teticha District to a total of 6,850 girls was estimated to be USD 9,470 – resulting in an average cost per vaccinated girl USD 1.38. The total cost of delivering HPV vaccination through the EPI integration model in Gomma District was USD 11,418 while the delivery cost per girl was USD 7.54. The total cost for delivering the HPV vaccine through the AH model in Sokoru District to a total of 1,284 girls was USD 18,347 for an incremental cost of USD 14.28 per girl.

**Table 4:** Total and incremental economic costs of three HPV vaccination models in sampled sites in USD.

| Cost category (USD) | MAC model | EPI model | AH model |
| --- | --- | --- | --- |
| **Capital** |  |  |  |
| Equipment | 862.3 | 415.1 | 415.1 |
| Vehicles | 1,739.2 | 821.4 | 818.8 |
| **Total capital costs** | **2,601.5** | **1,236.5** | **1,233.9** |
| **Recurrent** |  |  |  |
| Personnel | 3,573.3 | 4,114.5 | 8,553 |
| Supplies* | 1,842.5 | 991 | 387.6 |
| Vehicle operation and maintenance | 499.7 | 1,125 | 2,212.4 |
| Building operation and maintenance |  | 34.85 | 34.85 |
| Training | 953.3 | 1,812.8 | 1,629.8 |
| Workshops |  | 2,102.9 | 1,453.2 |
| Others (pads) |  |  | 2,842.5 |
| **Total recurrent costs** | **6,868.8** | **10,181.05** | **17,113.35** |
| **Total annual costs in sampled sites** | **9,470.3** | **11,417.55** | **18,347.25** |
| **Number of immunized girls in sampled sites\*\*\*** | **6,850** | **1,515** | **1,284** |
| **Incremental cost per vaccinated girl (USD)** | **1.38\*\*** | **7.54** | **14.28** |

\*Vaccine costs are excluded.
\*\*Not comparable due to opportunity costs and sustainability issues.
\*\*\*Number of immunized girls was estimated by summing the total number of girls vaccinated at each of the subset of sites included in the costing analysis for that specific arm from March 2025 (start of implementation) through July 2025.

## DISCUSSION

We found that two models for integrating HPV vaccination into routine healthcare delivery for girls ages 9 to 14 in Ethiopia performed well and were highly acceptable. More than 25,000 girls were vaccinated during the five-month project period, whereas no HPV vaccination occurred in comparison non-intervention areas during the same period. More than half of girls reached through these integrated models were not enrolled in school and likely were missed in the previous school-based campaign vaccination strategy, if eligible at that time. Qualitative research confirmed perceptions that community engagement, school involvement, and health facility linkages were central to successful implementation, while exit interview data indicated that clients served by both models had positive experiences and were satisfied with the care they received.

Community-centered outreach was a core component of both models. Integrating HPV vaccination into existing health and community systems allowed us to reach diverse adolescent populations, particularly out-of-school girls, while also ensuring coverage for in-school girls. These findings are in line with evidence from other settings, which illustrate how integrated, community-anchored delivery approaches can strengthen HPV vaccination outcomes. For example, a study conducted in Senegal on integration of HPV vaccination into routine vaccination showed that strong collaboration between health and education sectors, coupled with community engagement and mobilization through community members and local leaders, ensured sustained access and awareness, particularly among out-of-school girls [19]. The Smart Start platform, which the government of Ethiopia is currently scaling up nationally, facilitated the mapping and door-to-door identification of harder-to-reach unvaccinated girls. A major limitation of the MAC approach was the lack of a systematic process for locating eligible girls, which failed to fully reach populations like out-of-school girls. Thus, incorporating Smart Start’s established outreach platform into HPV vaccine delivery serves as a major advance.

Variations in performance and cost of the integration models can help guide strategies to scale up integrated vaccine delivery in Ethiopia and elsewhere. We found that facilities integrating HPV vaccination with AH services delivered fewer HPV vaccinations per month than did facilities integrating with the EPI program. These findings also held true for HPV vaccinations among out-of-school girls and those ages 9 to 12. Sensitivity analyses indicated that higher HPV vaccine delivery in the EPI arm may have been due in part to integration with the national measles and polio vaccination campaign that occurred in one EPI arm site and not in the other sites. Additionally, for girls ages 13 to 14, HPV vaccinations per month were similar across the two integration models. These results suggest age-specific differences in the intervention effect with a trend toward higher performance of the integrated AH model as girls age. This finding aligns with qualitative data suggesting that older girls may have been particularly interested in receiving menstrual health products. The AH model may therefore be particularly promising in areas where vaccine catch-up for older girls is the priority. Ongoing investments to strengthen adolescent-friendly health units within health centers should consider provision of HPV vaccination as a feasible and acceptable component of a broader adolescent health service offering. Thus, the preferred model may depend on a girl’s age, information and service needs, health status, and functionality of routine adolescent-focused health delivery platforms at the time of seeking care.

The cost per vaccinated girl for each of the three models assessed in our study falls within the range of costs for HPV vaccine delivery reported in other LMICs [20] and in Ethiopia [21], with the MAC model recording the lowest cost per girl vaccinated. The costs identified in our study also fall well below those reported recently from studies conducted in Zambia and Tanzania that showed costs of $28.1 and $23.34 per fully vaccinated girl [22,23]. Major cost-driving factors identified in our study were similar to those reported in previous studies, including start-up costs, service delivery, social mobilization, vaccine collection or distribution and storage at the health facility and administrative levels, and personnel costs [24–28]. While the prevailing periodic campaign-based approach has the lowest cost per vaccinated girl, likely in part due to the high volume of girls vaccinated per session, the higher cost of the EPI and AH models that incorporate ongoing outreach serve an important function in reaching girls who may never be reached through the lower cost MAC model.

Several unanticipated circumstances posed implementation and methodological challenges over the course of the study. For example, the unanticipated lack of AH service delivery at baseline made the set-up and delivery of the AH model more costly. Most critically, through formative research we learned that no ongoing HPV vaccination would occur during project implementation and the pre-implementation period in 2025. This reality, in conjunction with the short five-month duration of implementation, made it challenging to compare the effectiveness of the integrated models with the school-based campaign approach. We were able to strengthen the effectiveness analysis using monthly data and through adjustment for several potential confounders, including MAC performance and eligible population size immediately prior to start of the implementation. Nevertheless, residual confounding remains possible.

Several methodological limitations may constrain the generalizability of our findings, including the relatively low number of implementation sites. Furthermore, exit interviews were conducted in a purposive sample of implementation sites and therefore client perspectives may not be representative at the program level. The short five-month implementation period limited the assessment, making it difficult to evaluate long-term sustainability and program maintenance. Cost estimates from our study are also likely higher than they would be in longer-term ongoing implementation given the relatively limited duration of implementation in the study to offset major fixed costs, such as staff training, in estimates of costs per girl vaccinated for HPV. In addition, DHIS2 and costing data were dependent on the completeness and accuracy of routine records, which may have introduced reporting inconsistencies. Despite these limitations, the mixed-methods design provided a comprehensive, triangulated understanding of program performance and contextual challenges.

Strategies to scale-up integrated HPV vaccine delivery should be conditional on strengthening logistics, investing in comprehensive and practical training, institutionalizing supervision and documentation, and ensuring consistent provision of adolescent health services. Equally essential are sustained context-appropriate community engagement and social and behavior change interventions to address cultural concerns and misinformation. The system-level and community-level activities that were integrated into both the EPI and adolescent health arms in our study likely contributed to successful vaccine delivery across study arms. We recommend that a national strategy to scale-up integrated HPV vaccine delivery maintains the existing lower cost school-based campaign approach, while also incorporating integration with both routine immunization and adolescent health services to most efficiently reach adolescent girls who are seeking various services, including puberty education and menstrual health products and services. Integration with adolescent health services also provides an opportunity to revitalize national provision of adolescent-friendly health services to more broadly support adolescent health and wellbeing. An optimal mix of integration with the EPI program and adolescent health services should be determined based on regional HPV vaccination indicators, population demographics and characteristics of vaccine target populations, and costs of HPV vaccine delivery through each of the models.

## DATA AVAILABILITY STATEMENT

De-identified quantitative data and study instruments are available upon reasonable request to the authors. Due to the distinctive roles of some key informants interviewed, a subset of the qualitative data carries a higher risk of indirect identification and cannot be shared publicly to protect participants’ confidentiality; qualitative data deemed by the authors to carry low risk of indirect identification may be made available upon reasonable request.

## ACKNOWLEDGMENTS

The authors would like to thank all study team members from Population Services International and Addis Ababa University who contributed to the HPV-VISION research, M&E, and program implementation activities. We would also like to thank the study donors for their technical feedback on the study design and analytic approaches, which we believe greatly strengthened this research. We are grateful to the field teams involved in routine M&E and primary research data collection. Finally, we sincerely thank all study participants for contributing their time and valuable information.

## COMPETING INTERESTS

The authors declare no conflicts of interest.

## FUNDING

The HPV-VISION study was co-funded by Gavi: The Vaccine Alliance (GAVI100001742) and the Gates Foundation (INV-077693). The decision to publish was made solely by the authors.

## Notes

### Competing Interest Statement

The authors have declared no competing interest.

## REFERENCES

1 Jedy-Agba E, Joko WY, Liu B, et al. Trends in cervical cancer incidence in sub-Saharan Africa. Br J Cancer. 2020;123:148–54. doi: 10.1038/s41416-020-0831-9

2 World Health Organization. Cervical cancer. World Health Organization. 2025. https://www.who.int/news-room/fact-sheets/detail/cervical-cancer (accessed 16 March 2026)

3 Brisson M, Kim JJ, Canfell K, et al. Impact of HPV vaccination and cervical screening on cervical cancer elimination: a comparative modelling analysis in 78 low-income and lower-middle-income countries. The Lancet. 2020;395:575–90. doi: 10.1016/S0140-6736(20)30068-4

4 World Health Organization. Global strategy to accelerate the elimination of cervical cancer as a public health problem. Geneva: World Health Organization 2020.

5 Bruni L, Albero G, Serrano B, et al. Human Papillomavirus and Related Diseases in Ethiopia. 2023.

6 Federal Ministry of Health. Ethiopia National Expanded Program on Immunization: Comprehensive Multi-Year Plan (2021-2025). Addis Ababa, Ethiopia 2021.

7 Miazga W, Tatara T, Gujski M, et al. Global Guidelines and Trends in HPV Vaccination for Cervical Cancer Prevention. Med Sci Monit. 2025;31. doi: 10.12659/MSM.947173

8 World Health Organization. Human papillomavirus vaccines: WHO position paper (2022 update). Weekly Epidemiological Record. 2022;97:645–72.

9 Addisu D, Gebeyehu NA, Belachew YY. Knowledge, attitude, and uptake of human papillomavirus vaccine among adolescent schoolgirls in Ethiopia: a systematic review and meta-analysis. BMC Women’s Health. 2023;23:279. doi: 10.1186/s12905-023-02412-1

10 Woldehawaryat EG, Geremew AB, Asmamaw DB. Uptake of human papillomavirus vaccination and its associated factors among adolescents in Gambella town, Southwest, Ethiopia: a community-based cross-sectional study. BMJ Open. 2023;13:e068441. doi: 10.1136/bmjopen-2022-068441

11 Tiruye G, Sodo A, Tura AK, et al. Human papillomavirus vaccine uptake and associated factors among adolescent girls in Bona district, Sidama regional state, Ethiopia: a community-based study design. Front Public Health. 2025;13:1545171. doi: 10.3389/fpubh.2025.1545171

12 Gallagher KE, Howard N, Kabakama S, et al. Lessons learnt from human papillomavirus (HPV) vaccination in 45 low- and middle-income countries. PLoS ONE. 2017;12:e0177773. doi: 10.1371/journal.pone.0177773

13 Amponsah-Dacosta E, Blose N, Nkwinika VV, et al. Human Papillomavirus Vaccination in South Africa: Programmatic Challenges and Opportunities for Integration With Other Adolescent Health Services? Front Public Health. 2022;10:799984. doi: 10.3389/fpubh.2022.799984

14 Rosser EN, Wysong MD, Rosen JG, et al. HPV Vaccine Delivery Strategies to Reach Out-of-School Girls in Low- and Middle-Income Countries: A Narrative Review. Vaccines. 2025;13:433. doi: 10.3390/vaccines13050433

15 Rujumba J, Akugizibwe M, Basta NE, et al. Why don’t adolescent girls in a rural Uganda district initiate or complete routine 2-dose HPV vaccine series: Perspectives of adolescent girls, their caregivers, healthcare workers, community health workers and teachers. PLoS ONE. 2021;16:e0253735. doi: 10.1371/journal.pone.0253735

16 Jones N, Pincock K, Baird S, et al. Intersecting inequalities, gender and adolescent health in Ethiopia. Int J Equity Health. 2020;19:97. doi: 10.1186/s12939-020-01214-3

17 Ndiaye C, Kyesi F, Masupha T, et al. Integrating HPV vaccine service delivery with adolescent health programmes – Experiences and perspectives from selected countries in Africa. Vaccine. 2024;42:S45–8. doi: 10.1016/j.vaccine.2023.10.022

18 Akumbom AM, Lee JJ, Reynolds NR, et al. Cost and effectiveness of HPV vaccine delivery strategies: A systematic review. Preventive Medicine Reports. 2022;26:101734. doi: 10.1016/j.pmedr.2022.101734

19 Casey RM, Adrien N, Badiane O, et al. National introduction of HPV vaccination in Senegal— Successes challenges, and lessons learned. Vaccine. 2022;40:A10–6. doi: 10.1016/j.vaccine.2021.08.042

20 Slavkovsky R, Callen E, Pecenka C, et al. Costs of human papillomavirus vaccine delivery in low- and middle-income countries: A systematic review. Vaccine. 2024;42:1200–10. doi: 10.1016/j.vaccine.2024.01.094

21 Mvundura M, Bayeh A, Zelalem M, et al. Evaluating the cost and operational context for national human papillomavirus (HPV) vaccine delivery in three regions of Ethiopia. PLOS Glob Public Health. 2024;4:e0003357. doi: 10.1371/journal.pgph.0003357

22 Simuyemba MC, Chama-Chiliba CM, Chompolola A, et al. An evaluation of the cost of human papilloma virus (HPV) vaccine delivery in Zambia. BMC Infect Dis. 2024;24:369. doi: 10.1186/s12879-024-09222-2

23 Hsiao A, Struckmann V, Stephani V, et al. Costs of delivering human papillomavirus vaccination using a one-or two-dose strategy in Tanzania. Vaccine. 2023;41:372–9. doi: 10.1016/j.vaccine.2022.11.032

24 Levin CE, Van Minh H, Odaga J, et al. Delivery cost of human papillomavirus vaccination of young adolescent girls in Peru, Uganda and Viet Nam. Bull World Health Organ. 2013;91:585–92. doi: 10.2471/BLT.12.113837

25 Quentin W, Terris-Prestholt F, Changalucha J, et al. Costs of delivering human papillomavirus vaccination to schoolgirls in Mwanza Region, Tanzania. BMC Med. 2012;10:137. doi: 10.1186/1741-7015-10-137

26 Botwright S, Holroyd T, Nanda S, et al. Experiences of operational costs of HPV vaccine delivery strategies in Gavi-supported demonstration projects. PLoS ONE. 2017;12:e0182663. doi: 10.1371/journal.pone.0182663

27 Brennan T, Hidle A, Doshi RH, et al. Cost of human papillomavirus vaccine delivery in a single-age cohort, routine-based vaccination program in Senegal. Vaccine. 2022;40:A77–84. doi: 10.1016/j.vaccine.2021.11.057

28 Aldaba JG, Llave CL, Uy MaEV, et al. The cost of human papillomavirus vaccination delivery at the administrative and health facility levels in the Philippines. Vaccine: X. 2024;17:100459. doi: 10.1016/j.jvacx.2024.100459

