## Supplemental File for "Integrating HPV vaccination into adolescent health and routine immunization delivery platforms in rural and pastoralist Ethiopia: A mixed-methods implementation study of the HPV-VISION program"

**Figure S1. Vaccine delivery models and activities implemented across three study arms**

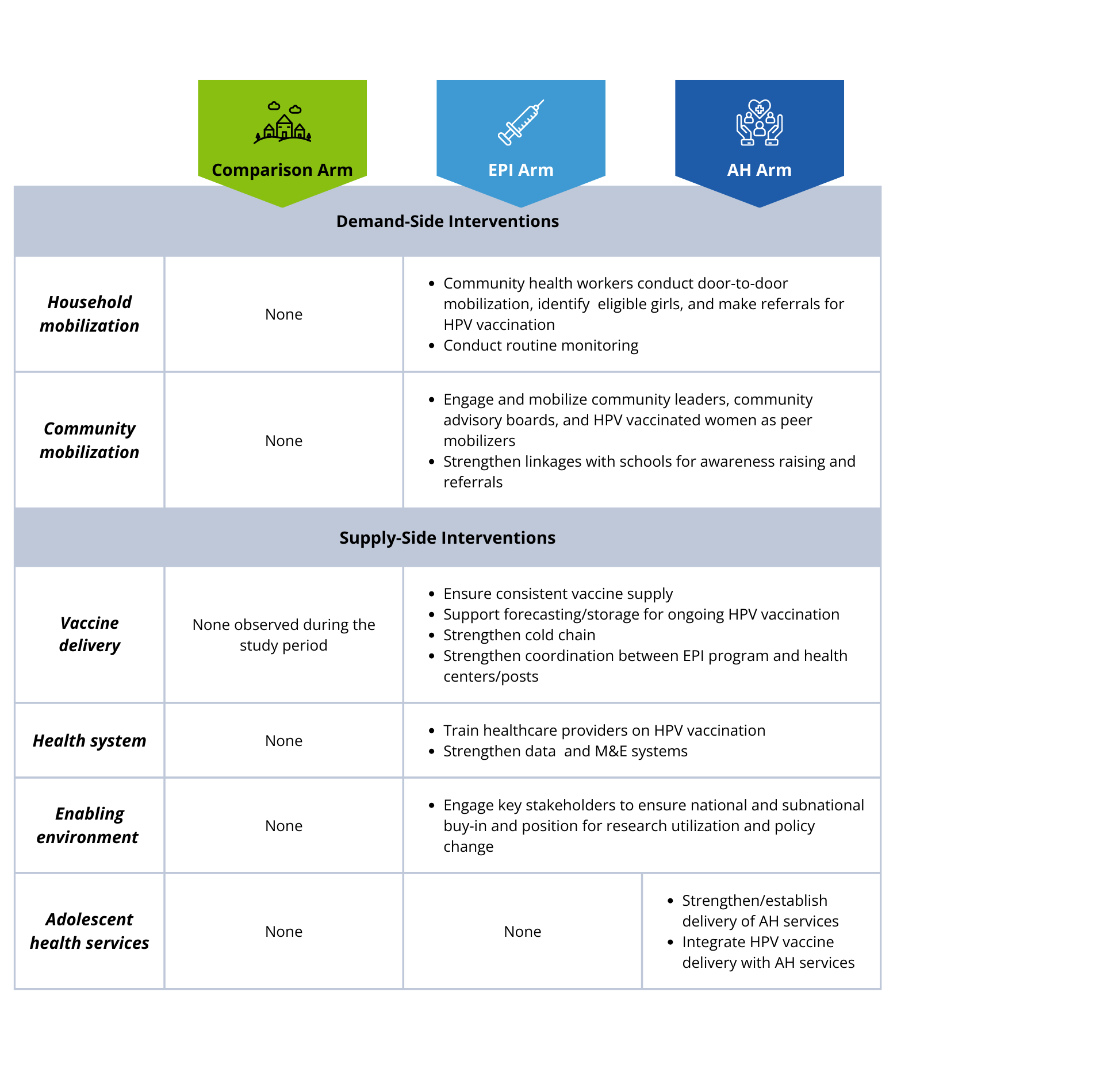

**Table S1. Facility-level HPV vaccinations delivered to girls ages 9 to 14 by study arm**

| **Measure** | **Comparison Arm**  n=131 facilities | **EPI Arm**  n=93 facilities | **AH Arm**  n=63 facilities |
| --- | --- | --- | --- |
| Total HPV vaccinations | 0 | 17,985 | 7,836 |
| Mean of total HPV vaccinations per facility | 0 | 193.4 | 124.4 |
| Median of HPV vaccinations per facility | 0 | 176 | 111 |
| *Mean of HPV vaccinations per facility per month (2025)* |  |  |  |
| March | 0 | 0.4 | 2.7 |
| April | 0 | 16.8 | 21.5 |
| May | 0 | 70.6 | 25.3 |
| June | 0 | 45.2 | 30.1 |
| July | 0 | 29.5 | 19.9 |
| August | 0 | 31.0 | 24.9 |
| Mean of total HPV vaccination of out-of-school girls per facility | 0 | 135.1 | 59.2 |
| *Mean of HPV vaccinations per facility by age group* |  |  |  |
| 9 | 0 | 49.5 | 24.1 |
| 10 | 0 | 36.8 | 23.1 |
| 11 | 0 | 32.2 | 16.1 |
| 12 | 0 | 28.9 | 17.2 |
| 13 | 0 | 24.3 | 19.2 |
| 14 | 0 | 21.7 | 24.7 |

**Figure S2. Secondary outcomes: Adjusted incidence rate ratios estimating the association between AH arm (vs. EPI arm) on HPV vaccinations of key subpopulations**

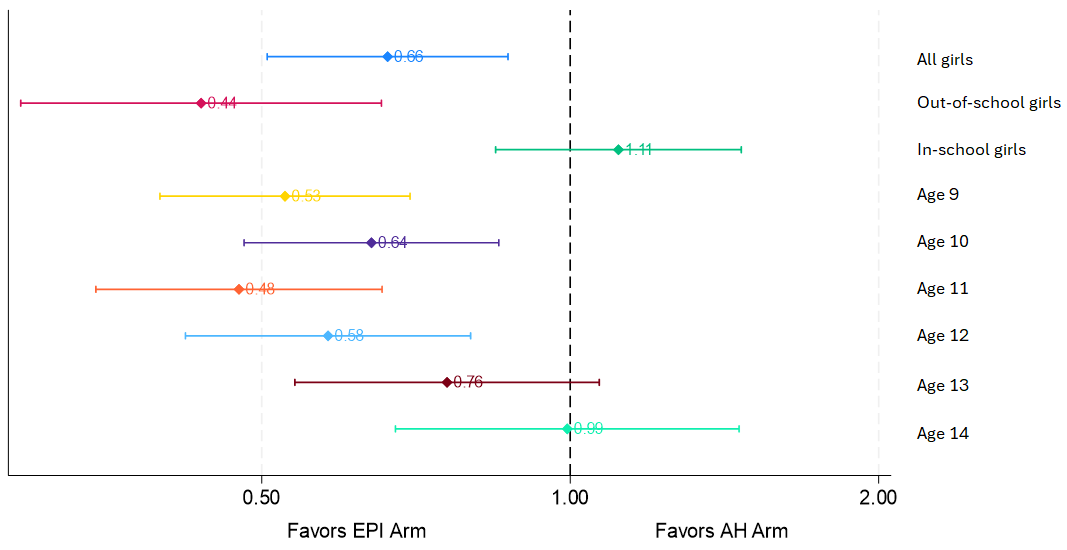

**Table S2. HPV vaccination delivery effectiveness: Secondary outcomes and sensitivity analyses**

| **Panel A. Secondary outcomes** | ***AH arm vs. EPI arm, IRR*** | ***p-value*** |
| --- | --- | --- |
| HPV vaccination counts of out-of-school girls | 0.44 | <0.001 |
| HPV vaccination counts of in-school girls | 1.11 | 0.446 |
| *HPV vaccination counts by age of girl* |  |  |
| 9 | 0.53 | <0.001 |
| 10 | 0.64 | 0.002 |
| 11 | 0.48 | <0.001 |
| 12 | 0.58 | 0.001 |
| 13 | 0.76 | 0.113 |
| 14 | 0.99 | 0.973 |
| **Panel B. Sensitivity analyses** | ***AH arm vs. EPI arm, IRR*** | ***p-value*** |
| Total HPV vaccinations, Poisson GEE | 0.47 | 0.001 |
| Total HPV vaccinations, excluding Awbare woreda | 0.79 | 0.136 |
| Total HPV vaccinations, excluding May | 0.71 | 0.071 |
| Notes: All models are adjusted for the following covariates: intervention, month, health facility type, MAC targets for in-school and out-of-school girls, and program targets for in- and out-of-school girls. IRRs represent the coefficient measuring the AH arm vs. the EPI arm (reference category). Each model includes 780 observations clustered within 156 health centers/posts. In Panel A, each row represents a separate negative binomial GEE model with a different secondary outcome. In Panel B, each row represents a model estimating the primary outcome (total HPV vaccination counts). The Poisson GEE includes 780 observations within 156 sites. The model excluding Awbare includes 515 observations within 103 sites. The model excluding May data includes 624 observations within 156 sites. | | |

**Table S3. HPV vaccination delivery effectiveness: Subgroup analyses**

| **Subgroup Analyses** | **Health Posts Only** | | **Standard Sites Only** | |
| --- | --- | --- | --- | --- |
|  | ***AH arm vs. EPI arm*** | | ***AH arm vs. EPI arm*** | |
| **Panel A. Fully adjusted models** | ***IRR*** | ***p-value*** | ***IRR*** | ***p-value*** |
| Total HPV vaccination counts | 0.89 | 0.248 | 0.64 | 0.005 |
| HPV vaccination counts of out-of-school girls | 0.73 | 0.032 | 0.42 | <0.001 |
| HPV vaccination counts of in-school girls | 1.17* | 0.238 | 1.13 | 0.421 |
| *HPV vaccination counts by age of girl* |  |  |  |  |
| 9 | 0.75 | 0.014 | 0.52 | <0.001 |
| 10 | 0.78 | 0.044 | 0.59 | 0.001 |
| 11 | 0.69 | 0.005 | 0.45 | <0.001 |
| 12 | 0.76 | 0.044 | 0.54 | 0.001 |
| 13 | 1.13 | 0.348 | 0.72 | 0.112 |
| 14 | 1.43 | 0.038 | 0.95 | 0.822 |
| Notes: Each row represents a separate negative binomial GEE model with a different secondary outcome. IRRs represent the coefficient measuring the AH arm vs. the EPI arm (reference category). Health post-only models include data from 130 health posts; standard site models include data from 143 facilities. All models include the following covariates: intervention, month, health facility type, MAC targets for in-school and out-of-school girls, and program targets for in- and out-of-school girls. | | | | |
| * Robust Poisson model used due to convergence issues | | |  |  |

**Table S4. Exploratory analysis assessing modification of the association intervention on HPV vaccination counts by facility type**

|  | ***IRR*** | ***p-value*** |
| --- | --- | --- |
| AH arm (*ref:* EPI arm) | 0.89 | 0.241 |
| Month (*ref:* April) |  |  |
| May | 2.29 | <0.001 |
| June | 1.79 | <0.001 |
| July | 1.26 | 0.055 |
| August | 1.39 | 0.001 |
| Health center (*ref*. health post) | 4.92 | <0.001 |
| **AH arm x health center (interaction term)** | **0.36** | **0.004** |
| *MAC campaign mapping and performance (November 2024)* | | |
| Total in-school eligible girls mapped | 1.00 | 0.079 |
| Total out-of-school eligible girls mapped | 1.00 | 0.007 |
| Number of in-school girls vaccinated | 1.00 | 0.079 |
| Number of out-of-school girls vaccinated | 1.00 | 0.028 |
| *HPV-VISION program mapping (March 2025)* |  |  |
| Total in-school eligible girls mapped | 1.00 | 0.047 |
| Total out-of-school eligible girls mapped | 1.01 | <0.001 |
| Intercept | 9.46 | <0.001 |
| Notes: Model includes 780 observations clustered within 156 health centers/posts. | | |

**Figure S3. Exploratory analysis of effect modification: Predicted HPV vaccination counts per facility, by intervention arm and facility type**

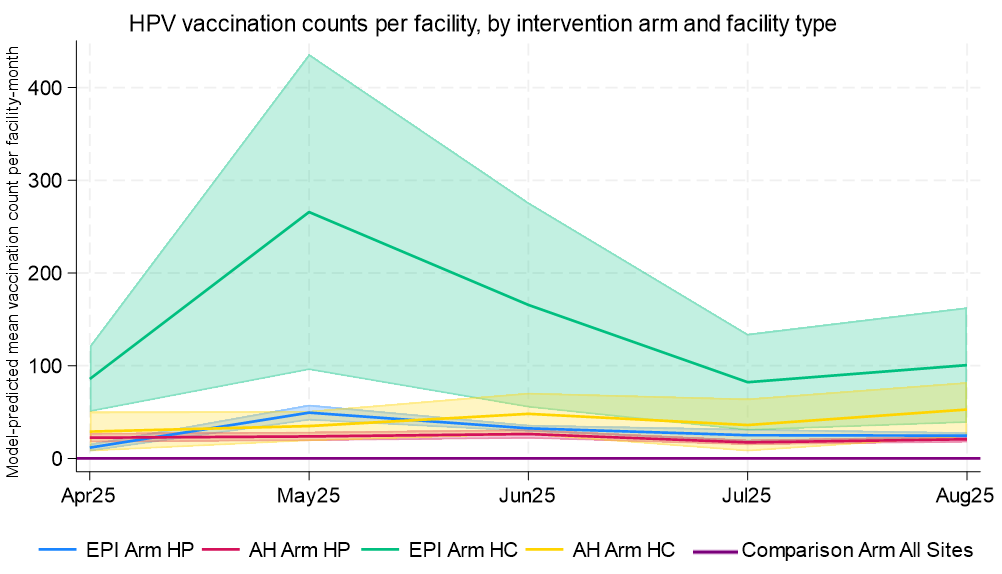
